# Number of plasma exchanges and outcome in myasthenic crisis

**DOI:** 10.1101/2022.01.31.22270211

**Authors:** Michael Hansen, Lee Neilson, Melanie Parikh, Bashar Katirji

## Abstract

**Background:** Plasma exchange is an effective therapy for myasthenic crisis (MC); yet the number of exchanges needed is unknown. We set out to examine the relationship between the number of plasma exchanges and clinical outcome in patients experiencing MC.

**Methods:** We retrospectively reviewed patient episodes with ICD 9 and ICD 10 codes for myasthenia gravis and myasthenia gravis exacerbation/crisis in patients admitted to a single center tertiary care referral center from July 2008 to July 2017. These episodes were screened for patients with impending myasthenia gravis crisis and manifest crisis who received plasmapheresis during their hospital course. We performed statistical analyses to determine if increased number of plasma exchanges improves the primary outcome (hospital length of stay), as well as the secondary outcome (disposition to home, skilled nursing facility, long term acute care hospital, or death).

**Results:** There is neither clinically observable nor statistically significant improvement in length of stay or disposition on discharge in patients who received six or greater sessions of plasmapheresis.

**Conclusions:** This study provides class IV evidence that extending the number of plasma exchanges beyond five does not correlate with decreased hospital length of stay or improved discharge disposition in patients experiencing myasthenic crisis.

## INTRODUCTION

Myasthenia gravis (MG) has an estimated prevalence of 150 to 250 cases per one million people, rendering it the most common neuromuscular junction disorder.^1^ Only 15-20% of patients with MG experience a myasthenic crisis (MC), usually during the first 2 years after diagnosis.^2,3^ Mortality rates drastically improved in the second half of the 20^th^ century due to the advent of effective long term immunosuppressive therapies which has been shown to reduce frequency of MC^4^, advances in effective invasive ventilation^5^, and in the emergence of rescue therapies specifically intravenous immunoglobulin (IVIg) and plasma exchange (PLEX).^6-8^ Patients with MC are often treated with PLEX or IVIg.^6^ Comorbidities, clinical status and availability often drive the selection of treatment between these modalities. PLEX may result in reduced days of mechanical ventilation and intensive care in MC when compared to IVIg,^6,9^ and expert consensus submit that PLEX works more quickly and is more effective.^10^ However, others found no differences in the efficacy of these two treatments.^11^

The therapeutic benefit of PLEX extends weeks beyond completion of a treatment course.^12,13^ However, it is an invasive procedure with multiple known complications including electrolyte derangements, metabolic alkalosis, catheter-associated thrombi, coagulation factor and immunoglobulin depletion, and transfusion-related complications.^14^ The most effective number of plasma exchanges in MC is not well established, though 4-5 PLEX sessions are common in clinical practice. In Guillain-Barre syndrome, a parallel acute neuromuscular disorder, two exchanges are indicated in mild disease and four in moderate and severe forms; additional exchanges add no benefit including in mechanically ventilated patients.^15^

Clinical data is lacking if extending PLEX beyond 5 sessions provides objective benefit. Yet clinicians may feel pressure to “do something^16^” such as continuing PLEX beyond five sessions if a patient hospitalized for MC is not responding to ongoing treatment, despite the inherent and significant risks of PLEX.^17^ We designed this study to examine the relationship between the number of PLEX sessions and clinical outcomes in patients with MC in our tertiary center.

## METHODS

### Study design and patient selection

We retrospectively identified patients, using ICD 9 and ICD 10 codes (358.0-358.01 and G70.0-70.3 respectively), with myasthenia gravis and myasthenia gravis exacerbation/crisis admitted to an urban tertiary care referral center, University Hospitals Cleveland Medical Center, from July 2008 to July 2017. Using these criteria, 1473 patients were identified (Figure 1). We screened these admissions for patients with impending and manifest MC^10^ *and* who received PLEX during their hospital course. One or more of the following confirmed the diagnosis of myasthenia gravis: elevated acetylcholine receptor (AChR) or muscle specific kinase antibodies (MuSK), abnormal repetitive nerve stimulation and/or abnormal jitter on single fiber EMG studies, or clinical diagnosis by a neuromuscular neurologist. We excluded patients with congenital myasthenia gravis, Lambert-Eaton myasthenic syndrome, cholinergic crisis or other myasthenic syndromes from the analysis. Two patients had more than one episode identified and we counted these separately as these patients were readmitted with a new trigger in-between episodes. We identified altogether 46 patients with 48 MC episodes that met the above criteria. The primary outcome was hospital length of stay, while the secondary outcome was disposition at discharge to home, skilled nursing facility, long-term acute care hospital, or death. Length of stay and disposition were determined to be evaluated prior to data collection as they were thought to be values most relevant to clinicians when determining the treatment course for patients suffering from myasthenic crisis.

**Figure 1:**
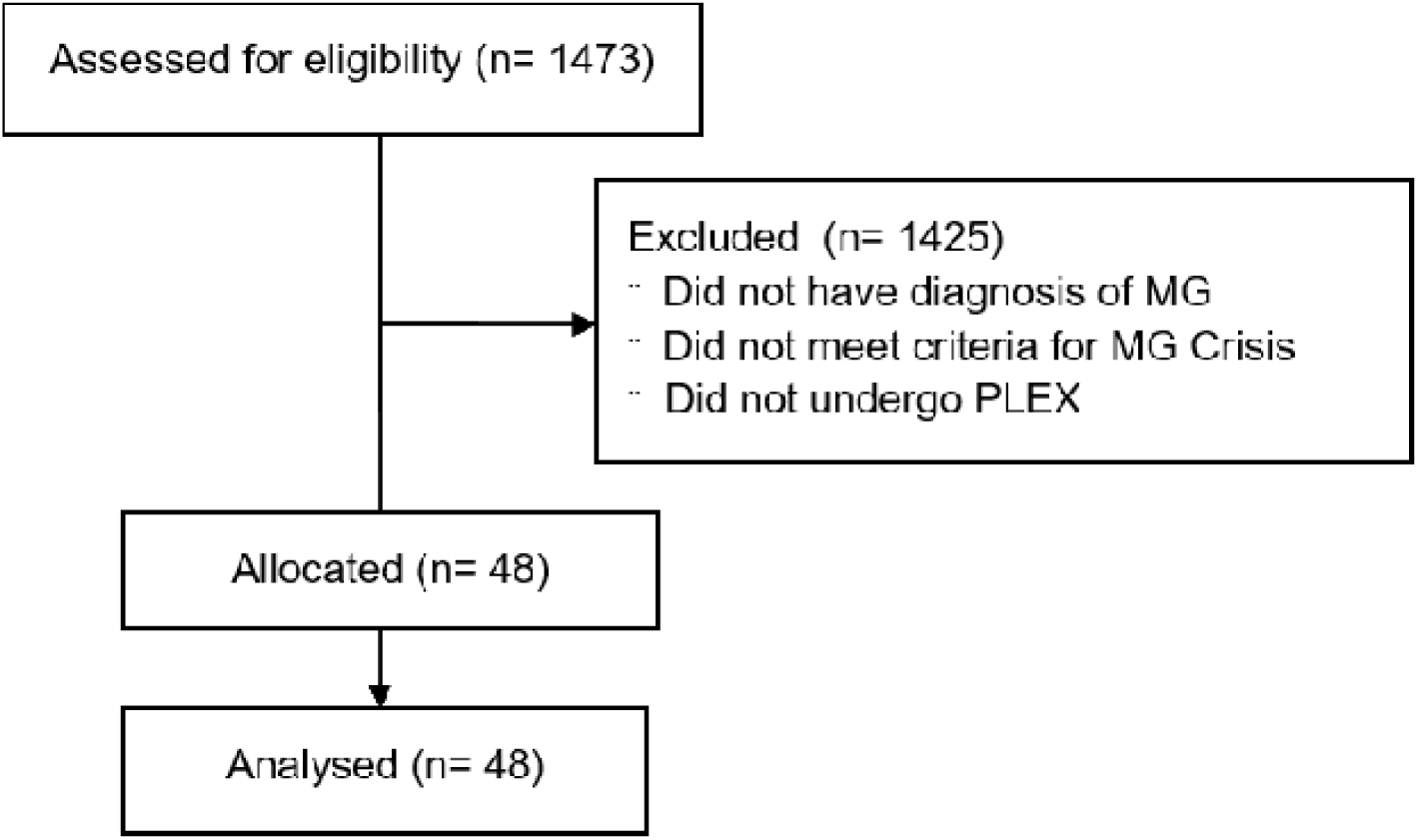
Flowchart mapping patient screening. Abbreviations: MG=myasthenia gravis; PLEX=plasma exchange

### Standard protocol approvals, registrations and patient consents

The University Hospitals Cleveland Medical Center Institutional Review Board and Ethics Committee approved this study. The data was collected retrospectively and anonymized, and patient consent was waived.

### Data acquisition

The data was acquired through chart review and searching of local electronic medical record databases. We collected baseline demographics including age and sex, as well as method of myasthenia gravis diagnosis on all patients. We recorded the characteristics of the patients’ myasthenia gravis including duration of disease, antibody titers, history of prior crises, thymoma (known or unknown) and history of thymectomy when available. Treatment data included use of steroids, steroid-sparing agents, acetylcholinesterase inhibitors, and intravenous immunoglobulins (IVIG). Co-morbidities including heart failure and chronic obstructive pulmonary disease were noted. We identified factors for crisis precipitation including treatment failure, infection, or contraindicated medication when available. Confounders arising during hospitalization including antibiotic use, plasmapheresis-related complications, line-related complications and electrolyte abnormalities were recorded. Data of the clinical course included the number of PLEX, number of days with invasive assisted ventilation, number of days in the hospital, and discharge disposition (defined as home, skilled nursing, long-term acute care facility, or hospice/death). When available, we recorded details about the plasmapheresis sessions including body volume exchanged, and replacement requirements.

### Statistics

PLEX sessions were first dichotomized to 5 sessions or fewer (n=39) and 6 sessions or more (n=9). For the primary outcome, hospital length of stay, we performed a Wilcoxon signed-rank test to evaluate non-parametric distributions with the significance level set at α = 0.05. For the secondary outcome, disposition at discharge, we performed a Chi-square test of home, skilled nursing facility, long-term care facility, and hospice/death. A large confounding variable is that more PLEX sessions are often deemed necessary in patients with more severe crises. To address this concern, we identified patients who remained on assisted invasive ventilation at the time of their fifth exchange, the time when clinicians would often need to opt for additional exchanges. Statistical analysis was repeated for the subgroup of 5 sessions of PLEX (n=9) vs 6 or more sessions of PLEX (n=5). The R Statistical software, version 3.6 (R Foundation for Statistical Computing, Vienna, Austria) was used for statistical analyses.

### Data availability

Raw data will be shared if requested.

## RESULTS

### Baseline characteristics

We identified 48 episodes in 46 patients who experienced MC and received PLEX over a 9 year period at our institution. Baseline characteristics are identified in table 1. The median age of the patients at the time of crisis was 65.5 years (range 21-90 years of age); 50% of patients were women. In 11 patients (24%), MC was their initial presentation of the disease. Overall, 38 (83%) patients were seropositive. Specific antibody results were available in 33 patients, with 28 of 46 having elevated AChR antibody (61%) and five (11%) elevated MuSK antibody. Five patients (11%) were noted as “seropositive” in the chart but the specific antibody elevation was not available for review. Eight (17%) seronegative patients were diagnosed based on electrodiagnostic studies or clinical assessment by neuromuscular specialist. We identified 39 patients who had thoracic imaging prior to or during the MC under investigation; thymoma was identified in five (11%) patients. Thymectomy was done prior to the presenting MC in11 (24%) patients. Comorbidities included one patient (2%) patient with known COPD, four (9%) with heart failure, and 12 (26%) with other co-morbidities (asthma, anemia, arrhythmia, deep vein thrombosis, malignancy, obstructive sleep apnea).

**Table 1.** Baseline characteristics of 46 myasthenia gravis patients hospitalized with 48 myasthenic crises. Abbreviations: No.=number; y=years; RNS=repetitive nerve stimulation; SFEMG=single fiber electromyography; AChR=acetylcholine receptor; MuSK=muscle-specific receptor tyrosine kinase; DVT=deep venous thrombosis; OSA=obstructive sleep apnea; IVIG=intravenous immunoglobulin

|  | No. (%) or median (range) |
| --- | --- |
| **Baseline patient characteristics** |  |
| No. of patients | 46 |
| No. of myasthenic crisis episodes | 48 |
| Female | 23 (50) |
| Age, y | 65.5 (21-90) |
| Myasthenic crisis as first presentation | 11 (24) |
| Primary method of diagnosis |  |
| Serology | 38 (83) |
| RNS, SFEMG or clinical exam | 5 (11) |
| Clinical exam | 3 (7) |
| Specific antibody status |  |
| AChR | 28 (61) |
| MuSK | 5 (11) |
| Seronegative | 8 (17) |
| Unknown | 5 (11) |
| Thymus status |  |
| Thymoma | 5 (11) |
| Known thymectomy | 11 (24) |
| Chronic obstructive pulmonary disease | 1 (2) |
| Heart failure | 4 (9) |
| Other co-morbidities (asthma, anemia, arrhythmia, DVT, malignancy, OSA) | 12 (26) |
| **Myasthenic crisis characteristics** |  |
| Treatment regimen at the onset of crisis |  |
| Pyridostigmine | 37 (77) |
| Prednisone | 29 (60) |
| Steroid-sparing agents (azathioprine, mycophenolate mofetil) | 11 (23) |
| Outpatient maintenance IVIG | 5 (10) |
| Trigger for myasthenic crisis |  |
| Infection | 9 (19) |
| Changes in myasthenia treatment regimen | 5 (10) |
| Checkpoint inhibitor (ipilimumab, nivolumab) | 2 (4) |
| Contraindicated medication | 2 (4) |
| Post-operative state | 1 (2) |
| Medication non-adherence | 1 (2) |
| In-hospital IVIG treatment for myasthenic crisis | 17 (35) |
| Plasmapheresis for myasthenic crisis |  |
| Sessions | 5 (3-11) |
| $\leq 5$ | 39 (81) |
| $\geq 6$ | 9 (19) |

### Myasthenic crisis characteristics

At the time of hospital admission for MC, 37 (77%) patients were taking pyridostigmine, 29 (60%) prednisone, 11 (23%) steroid-sparing agents (azathioprine or mycophenolate mofetil), and 5 (10%) maintenance pulse IVIG. In those patients not on prednisone at admission, hospital records were not sufficient to determine consistently when prednisone was started in the course of the myasthenic crisis. A reason for crisis was suspected in 20 (42%) episodes. Of those, nine were suspected to be due to infection, five due to changes in MG treatment regimen, two due to use of immune checkpoint inhibitors (ipilimumab/nivolumab), two due to use of medications known to be contraindicated in MG, one due to medication non-adherence and one in the perioperative setting. Patients received IVIG while hospitalized for MC in 17 of 48 (35%) of episodes. Eleven of 39 (28%) who received ≤5 exchanges received IVIG compared to six of nine (66%) in patients who received ≥6 exchanges. Using a Chi-squared analysis, we found the difference to be statistically significant (p=0.030). In patients who required mechanical ventilation at the time of the fifth exchange, five of nine (56%) who received five exchanges received IVIG compared to four of five (80%) who received ≥6 exchanges. The difference was not significantly significant (p=0.36).

### Plasma exchange characteristics

The number of PLEX sessions varied from 3 to 11 sessions with the most frequent regimen being five exchanges in 26 of 48 (54%) episodes. We made every attempt to identify the body volume exchanged as well as fluid replacement characteristics for each PLEX session; however, these values were inconsistent and thus, deemed too unreliable to perform further statistical analysis. Most sessions exchanged one body volume; however, there were a number of sessions identified that exchanged 1.5 body volumes for the first exchange.

### Outcomes

Patients who received ≤5 plasma exchanges had a significantly lower hospital length of stay (LOS) vs those receiving ≥6 plasma exchanges (median value 10 days vs 29 days, p=0.0041; figure 2A). The difference in disposition between the two groups was also statically significant (p=0.020; figure 3A). Comorbidities occurred in similar rates between the two groups. In an attempt to compare only those with the most severe crises which likely triggered the increased number of exchanges, we identified 14 patients with MC episodes that remained mechanically ventilated after their fifth PLEX. Nine of these patients received five plasma exchanges and five patients received ≥6 plasma exchanges. The mean hospital LOS for patients receiving five sessions was significantly less compared to patients receiving six or greater sessions (median value 23 vs 38 days, p=0.023; figure 2B). Disposition at discharge was not significantly different (p=0.18; figure 3B). Comorbidities occurred in similar rates between the two groups.

**Figure 2:**
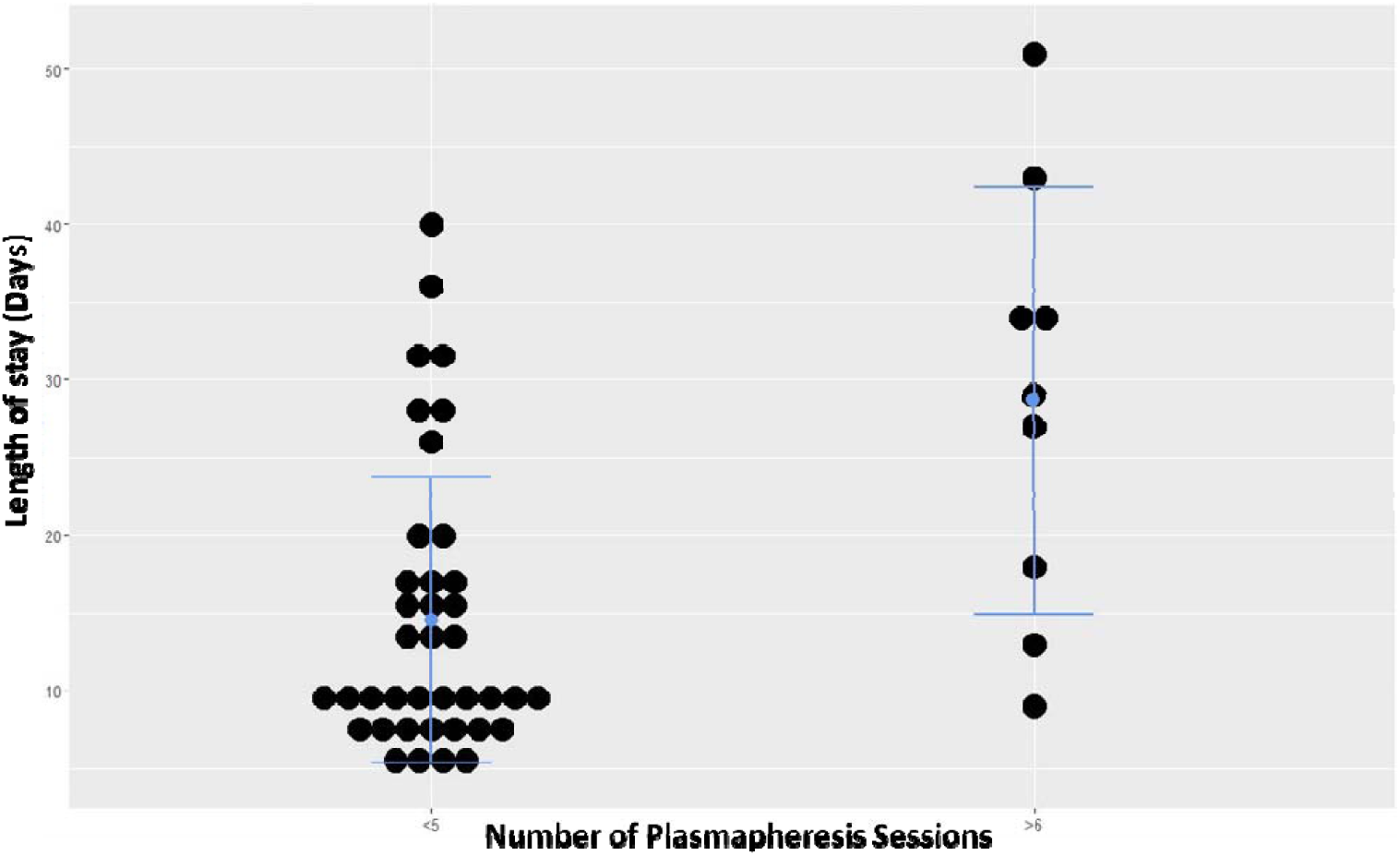
Boxplots comparing length of stay between (A) ≤5 exchange group (n=39) to ≥6 exchange group (n=9) and (B) 5 exchange group (n=9) to ≥6 exchange group (n=5) in patients who required mechanical ventilation at the time of their fifth exchange. Box shows first quartile, second quartile (median) and third quartile with whiskers showing the minimum and maximum values. Quartiles overlap in ≥6 exchange group shown in B. Gray dots show outliers. Abbreviations: PLEX=plasma exchange

**Figure 3:**
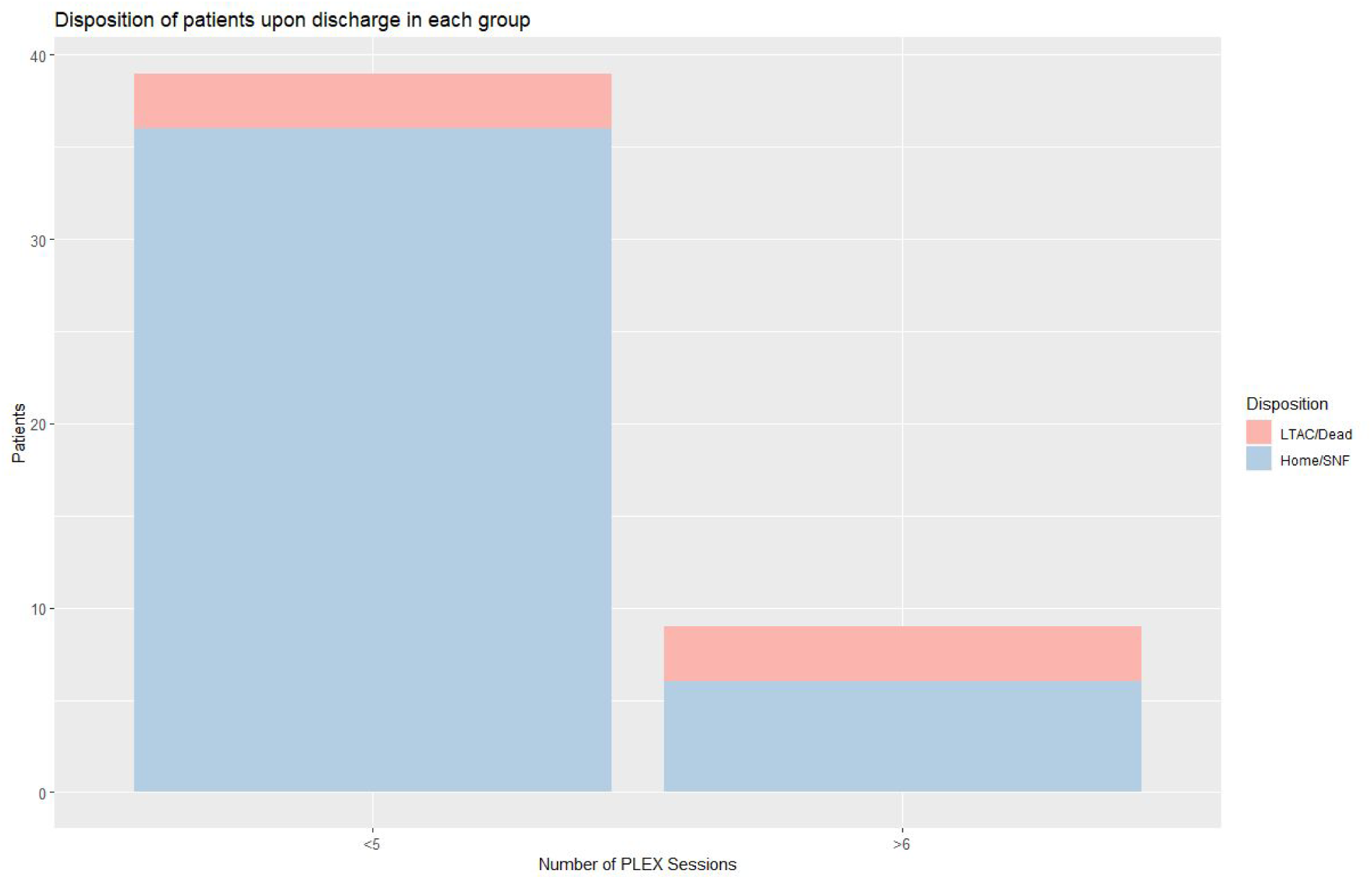
Bar graphs reporting disposition between (A) ≤5 exchange group (n=39) to ≥6 exchange group (n=9) and (B) 5 exchange group (n=9) to ≥6 exchange group (n=5) in patients who required mechanical ventilation at the time of their fifth exchange. Abbreviations: PLEX=plasma exchange; SNF=skilled nursing facility; LTAC=long term acute care hospital

## DISCUSSION

Polyclonal IgG antibodies are pathogenic in MG through activation of complement cascade at the post-synaptic junction, enhanced endocytosis of acetylcholine receptors, and direct blocking of ACh binding site.^18^ Given that PLEX removes these autoantibodies and is an effective^12^, and possibly preferred^19^, therapy for MC, it is essential to understand if there should be variability in the number of sessions administered.^20^ There is evidence that increasing PLEX between 3 and 5 sessions results in a reduced rate of MG relapse during the ensuing 50 months.^21^ A typical course of treatment in MC is often five PLEX sessions.^12^ Data suggest that PLEX removes about 70% of intravascular IgG antibody per body volume exchanged and plateaus by five sessions.^22,23^ As the number of exchange sessions increases, the yield of antibody removal decreases. These foundational facts of the mechanism of pathogenesis of IgG antibodies in MG provide a strong physiological argument that extension of PLEX beyond five sessions for MC has significantly diminishing benefits while exposing the patient to a greater risk for complications. However, this approach may be too simplistic, as it ignores extravascular sources and repletion of IgG levels. Extending PLEX in MC may have clinical efficacy by influencing the re-synthesis or even rebound production of the autoantibodies since the physiologic effects of PLEX are not limited to IgG removal.^24-27^

This study represents the largest single center study addressing the number of PLEX sessions in MC. Our patient population is comparable in several characteristics of MC to a recently published, large multi-center German cohort.^6^ MC was the first manifestation of their disease in 24% of our patients compared with 21%. Infection was the most likely identified cause for MC in both studies; however, our overall identification of suspected precipitating factor of MC was lower than in prior studies.^3^ The average LOS in the hospital in our study was significantly different (17 days versus 31 days), a wide gap likely representing selection of different populations (those who received PLEX only versus all MC mechanical ventilation) and possibly for inherent differences in healthcare delivery between the United States and Germany.

When comparing our patients who received ≤5 sessions of PLEX to ≥6 sessions of PLEX, hospital LOS and outcome did not improve. In fact, hospital LOS was significantly longer in those patients who received more than five PLEX sessions, likely reflecting that patients with more severe crises are more likely to undergo more exchanges. To address this, we identified those patients who remained mechanically ventilated after their fifth session of PLEX to attempt to compare only those with the most severe crises. The results did not differ; those who received ≥6 sessions of PLEX did not have shorter hospital LOS or improved outcomes. Both groups who remained intubated at the time of the fifth session of PLEX received IVIG at some point during the hospitalization at high rates. The difference was not found to be significantly different, however due to the small numbers of patients, it is difficult to comment further regarding the relationship of IVIG and clinical outcome in our dataset. Overall, our data does not support extending PLEX beyond 5 sessions for MC. The anecdotal clinical improvement sometimes seen during the period of extended exchanges likely reflects, in part, the effects of repair of the neuromuscular junction.

Our study has several limitations. *First*, our data is retrospective. Prospective, controlled data would be ideal; however, given the rarity and variability of MC, patient recruitment would be extremely challenging as clinicians may balk at relinquishing the ability to tailor treatments to individual MC. Such a study would also require a coordinated, multicenter study complicated by variability in treatment regimen among different institutions as previously noted in the literature.^9^ *Second*, the severity of symptoms in patients using a validated scale, such as the quantitative myasthenia gravis score, was not included in the analysis since documentation was not sufficient to reliably determine values. *Third*, our study is limited to one tertiary care institution rendering our patient population and management schemes possibly not representative for all patients suffering from MC.

In conclusion, our study provides level IV evidence that increasing plasma exchanges beyond five sessions does not improve outcome as measured by decreased LOS or improved disposition. Additionally, ≥6 exchanges did not correlate with decreased LOS or improved disposition in refractory cases of MC.

## Data Availability

Raw data will be shared if requested.

## Appendix 1: Authors

| Name | Location | Contribution |
| --- | --- | --- |
| Michael Hansen, MD | University Hospitals<br>Cleveland Medical Center and Case Western Reserve University,<br>Cleveland OH, | Design and conception of study; data acquisition; data analysis; drafted the manuscript for intellectual content |
| Lee Neilson, MD | University Hospitals<br>Cleveland Medical Center and Case Western Reserve University,<br>Cleveland, OH | Design and conception of study; data acquisition; drafted the manuscript for intellectual content |
| Melanie Parikh, MD | Case Western Reserve University,<br>Cleveland, OH | Data acquisition; data analysis; revised the manuscript for intellectual content |
| Bashar Katirji, MD | University Hospitals<br>Cleveland Medical Center and Case Western Reserve University,<br>Cleveland, OH | Design and conception of study; data analysis; drafted the manuscript for intellectual content |

